# SARS-CoV-2 Antibody Responses Correlate with Resolution of RNAemia But Are Short-Lived in Patients with Mild Illness

**DOI:** 10.1101/2020.08.15.20175794

**Authors:** Katharina Röltgen, Oliver F. Wirz, Bryan A. Stevens, Abigail E. Powell, Catherine A. Hogan, Javaria Najeeb, Molly Hunter, Malaya K. Sahoo, ChunHong Huang, Fumiko Yamamoto, Justin Manalac, Ana R. Otrelo-Cardoso, Tho D. Pham, Arjun Rustagi, Angela J. Rogers, Nigam H. Shah, Catherine A. Blish, Jennifer R. Cochran, Kari C. Nadeau, Theodore S. Jardetzky, James L. Zehnder, Taia T. Wang, Peter S. Kim, Saurabh Gombar, Robert Tibshiran, Benjamin A. Pinsky, Scott D. Boyd

**Affiliations:** Department of Pathology, Stanford University School of Medicine, Stanford, CA, USA.; Stanford ChEM-H and Department of Biochemistry, Stanford University School of Medicine, Stanford, CA, USA.; Department of Structural Biology, Stanford University, Stanford, USA.; ATUM, Newark, CA, USA; Stanford Blood Center, Palo Alto, CA, USA.; Department of Medicine, Division of Infectious Diseases and Geographic Medicine, Stanford University, Stanford, CA, USA.; Department of Medicine, Division of Pulmonary, Allergy and Critical Care Medicine, Stanford University, Stanford, CA, USA.; Stanford Center for Biomedical Informatics Research, Stanford University, Stanford, California, USA; Chan Zuckerberg Biohub, San Francisco, CA, USA.; Department of Bioengineering, Stanford University, Stanford, CA, USA.; Sean N. Parker Center for Allergy and Asthma Research, Stanford, CA, USA.; Department of Microbiology and Immunology, Stanford University, Stanford, CA, USA.; Department of Biomedical Data Sciences, Stanford University, Stanford, CA, USA.; Department of Statistics, Stanford University, Stanford, CA, USA.

## Abstract

SARS-CoV-2-specific antibodies, particularly those preventing viral spike receptor binding domain (RBD) interaction with host angiotensin-converting enzyme 2 (ACE2) receptor, could offer protective immunity, and may affect clinical outcomes of COVID-19 patients. We analyzed 625 serial plasma samples from 40 hospitalized COVID-19 patients and 170 SARS-CoV-2-infected outpatients and asymptomatic individuals. Severely ill patients developed significantly higher SARS-CoV-2-specific antibody responses than outpatients and asymptomatic individuals. The development of plasma antibodies was correlated with decreases in viral RNAemia, consistent with potential humoral immune clearance of virus. Using a novel competition ELISA, we detected antibodies blocking RBD-ACE2 interactions in 68% of inpatients and 40% of outpatients tested. Cross-reactive antibodies recognizing SARS-CoV RBD were found almost exclusively in hospitalized patients. Outpatient and asymptomatic individuals’ serological responses to SARS-CoV-2 decreased within 2 months, suggesting that humoral protection may be short-lived.

## Introduction

A novel coronavirus first described in Wuhan, China in December 2019 *(1)*, has led to a coronavirus disease (COVID-19) pandemic and a global economic shutdown amid unprecedented social distancing measures. The clinical spectrum of COVID-19 ranges from asymptomatic infection and mild upper respiratory tract illness in the majority of patients, to severe viral pneumonia with respiratory failure, multiorgan failure, and death *(2–4)*. Initial indications are that older adults and people with serious underlying health conditions are at greatest risk for severe illness *(5–7)*. Host immune system responses may be one of the most important determinants for disease progression and outcome.

The virus causing COVID-19 belongs to the *Sarbecovirus* subgenus (genus *Betacoronavirus*) together with the *Severe acute respiratory syndrome-related coronavirus* (SARS-CoV) and has been designated SARS-CoV-2 *(8)*. Coronaviruses contain four structural proteins, including spike, envelope, membrane, and nucleocapsid (N) proteins. The spike surface glycoprotein, which contains RBD, plays a major role in viral attachment, fusion of viral and host membranes, and entry of the virus into host cells and is a determinant of host range and tissue tropism *(9)*. SARS-CoV-2 RBD binds strongly to human ACE2 receptors *(1, 10)*, and is likely an important target for virus neutralizing antibodies. The highly immunogenic spike protein or RBD alone are therefore targets of interest for the development of serological and neutralization assays. Serological surveillance is of critical public health importance to monitor SARS-CoV-2 infection prevalence, death rate, and the eventual development of herd immunity, as well as to identify potential donors of convalescent plasma for therapeutic use *(11)*. Virus-specific antibodies develop within 1 to 2 weeks after COVID-19 symptom onset and can also aid in diagnosis of infections in those for whom reverse transcription polymerase chain reaction (RT-PCR) testing of respiratory tract specimens for viral RNA is negative *(12)*.

Building on initial reports from China and Europe, there is an urgent need for better understanding of the relationships between virus-specific antibody responses, SARS-CoV-2 persistence in the host, and the clinical course and outcome for patients *(13–15)*.

To address this need, we performed a comprehensive analysis of antibodies raised to the SARS-CoV-2 spike RBD and S1 domains, and the N protein in 494 plasma samples from 84 outpatients, 25 non-ICU (intensive care unit) inpatients, and 15 ICU patients, in the Stanford Healthcare system. More limited serological testing provided data for IgM and IgG responses to RBD in an additional 86 outpatients and asymptomatic individuals. SARS-CoV-2 IgM, IgG, and IgA isotype responses show distinct time courses and variation with antigen type and are strongly associated with the clinical severity of infection. Increasing quantities of antibodies in the blood, which showed increasing ACE2 receptor blocking activity, were negatively correlated with viral RNAemia. Outpatients and asymptomatic individuals have particularly short-lived plasma antibody responses, decreasing after the first month of documented infection.

## Results

### Study design and patient demographics

210 individuals with positive SARS-CoV-2 real-time RT-PCR (rRT-PCR) nasopharyngeal swab tests were included in the study (**fig. S1**). Patients with symptoms of COVID-19 either reported to Stanford Healthcare-associated clinical sites or were identified as having SARS-CoV-2 infection through occupational health screening with rRT-PCR and serology testing at Stanford Clinical Laboratories for anti-SARS-CoV-2 RBD IgM and IgG antibodies.

In total, 40 inpatients were included, of whom 15 required ICU care, and three died of COVID-19. 170 outpatients or asymptomatic individuals were included in the study. For all 40 inpatients, and for 84 of the outpatients, remnant plasma samples from diagnostic testing were available for a detailed research analysis of antibody responses to SARS-CoV-2 and SARS-CoV (**Fig. 1**). Stanford Clinical Lab serologic testing for IgM and IgG antibodies to SARS-CoV-2 RBD antigen was performed on samples from only 149 of the 170 outpatient or asymptomatic individuals, because this testing was not available at the time that the remaining 21 individuals underwent rRT-PCR testing.

**Fig. 1.**
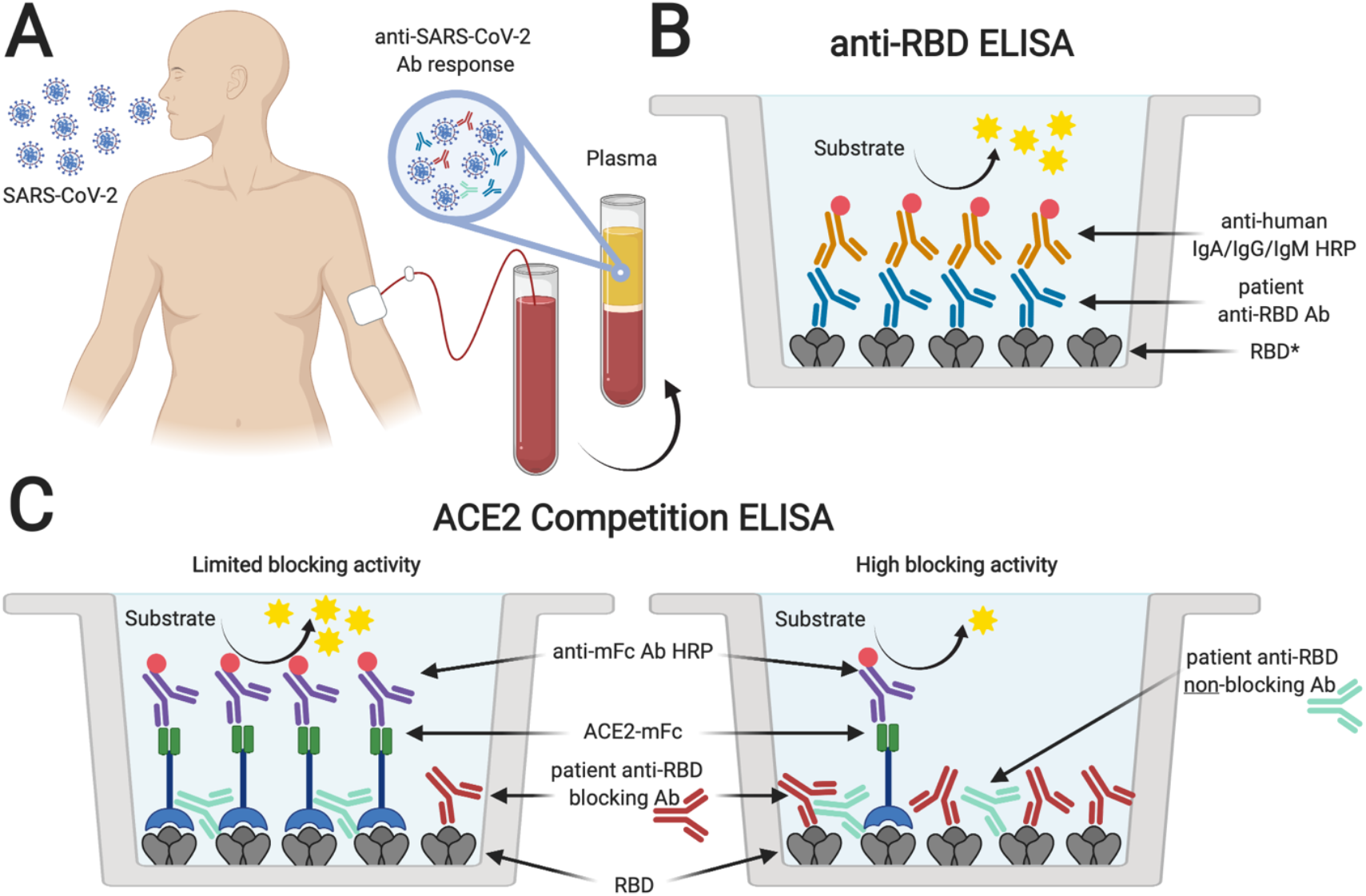
Serological testing of plasma from SARS-CoV-2 PCR+ individuals. Plasma samples from SARS-CoV-2 rRT-PCR-positive individuals (**A**) were analyzed for the presence of antibodies binding to SARS-CoV-2 spike RBD (**B**). *Plasma was also tested for antibodies specific for SARS-CoV-2 S1 and N protein, and SARS-CoV RBD. In addition, samples were tested for antibodies blocking the interaction of ACE2 and RBD in an ACE2 competition ELISA (**C**). Absence or limited presence of anti-RBD antibodies resulted in ACE2 binding to RBD and increased ELISA signals, whereas the presence of blocking antibodies prevented binding of ACE2, resulting in lower ELISA signals (created with biorender.com).

The median age of the 84 outpatients studied in more detail was 40 years (inter-quartile range (IQR) 31–56) with 32 men and 52 women. The median age of the 40 inpatients was 57 years (IQR 42–70) with 19 men and 21 women. Demographic and clinical characteristics of inpatients are presented in **Table 1**. The median number of plasma samples collected from ICU and admitted non-ICU patients respectively, were 11 (IQR 9–28) and 5 (IQR 3–7). Of the 84 outpatients, 8 had 2 time points, two others had 3 and 4 time points, and the remainder had a single time point sampled. The date of symptom onset was available for 38 of the 40 inpatients and all of the 84 outpatients. A total of 494 samples from the 124 inpatients and outpatients were available for detailed serological testing.

**Table 1.** Patient Demographic and Clinical Characteristics.

| **Characteristics** |  | **Admitted, non-ICU**<br>**(*n*=25)** | **Admitted, ICU**<br>**(*n*=15)** |
| --- | --- | --- | --- |
| Age (median (IQR)) |  | 55 (42-66) | 58 (43-71) |
| Sex (%) | Female | 14 (56.0) | 7 (46.7) |
|  | Male | 11 (44.0) | 8 (53.3) |
| Comorbidities<br>(% present) | Obesity | 9 (37.5) | 10 (66.7) |
|  | Hypertension | 5 (20.8) | 7 (46.7) |
|  | DM2 | 4 (16.7) | 4 (26.7) |
|  | Asthma | 3 (12.5) | 2 (13.3) |
| Symptoms on presentation<br>(% present) | Cough | 22 (88.0) | 15 (100) |
|  | Fever | 19 (76.0) | 11 (73.3) |
|  | SOB | 18 (72.0) | 14 (93.3) |
|  | Myalgia | 12 (50.0) | 8 (53.3) |
|  | GI | 14 (56.0) | 6 (40.0) |
|  | Fatigue | 14 (58.3) | 7 (46.7) |
|  | Chills | 7 (29.2) | 8 (53.3) |
|  | Headache | 5 (20.8) | 5 (33.3) |
| Mechanical ventilation (%) |  | 0 | 9 (60.0) |
| 30-day all-cause mortality (%) |  | 0 | 3 (20.0) |
| Length of hospital stay<br>(median days (IQR)) |  | 8.5 (4-11) | 10 (4-19) |

ICU, intensive care unit; IQR, inter quartile range; DM2, Diabetes mellitus type 2; SOB, shortness of breath; GI, gastrointestinal.

### Anti-RBD antibody responses and duration are associated with disease status

Low antibody responses in asymptomatic and mildly infected patients have been reported for other coronavirus infections, such as MERS-CoV *(16–18)*. In 396 plasma samples from 40 COVID-19 inpatients in this study, the detection rate of antibodies binding to the viral spike RBD at 1, 2, 3, and 4 weeks after symptom onset was 27%, 70%, 91% and 98% (IgM), 30%,73%, 94%, and 98% (IgG), and 27%, 67%, 90%, and 98% (IgA), respectively. All inpatient samples at five weeks post-onset of symptoms were positive for anti-RBD IgM, IgG, and IgA. Positivity rates for anti-RBD IgM and IgA began to drop thereafter, while anti-RBD IgG levels persisted over a longer time period (**Fig. 2A, table S1**). In contrast to the inpatient data, serological testing of plasma from 149 outpatients and asymptomatic individuals at Stanford Clinical Laboratories showed lower rates of positivity for anti-SARS-CoV-2 RBD IgM and IgG, with a peak of 74% positive for IgM and 87% for IgG in week 4 after the first positive SARSCoV-2 rRT-PCR test. Sero-positivity rates of the plasma samples dropped rapidly after this peak (**Fig. 2B, table S2**). Taking all timepoints at least 20 days after the first SARS-CoV-2 rRT-PCR positive test for outpatients and asymptomatic individuals, or after symptom onset for inpatients, the slope for the decrease in anti-RBD IgG titers was greater (p<0.001) for outpatients and asymptomatic individuals than for inpatients (slopes of –3.17 and –1.12, respectively). Measured antibody levels were significantly lower for outpatients with a high diagnostic SARS-CoV-2 rRT-PCR cycle threshold (Ct), indicating low levels of viral RNA in nasopharyngeal swab samples, as compared to those with a low Ct and high levels of viral RNA (**Fig. 2C**). The time course for IgM and IgG positivity in 84 outpatients with known date of symptom onset confirmed the results seen for the larger dataset of samples from 149 outpatients and asymptomatic individuals plotted as a function of the date of the first positive rRT-PCR test.

Overall fewer outpatients as compared to inpatients developed anti-RBD IgA titers (**Fig. 2D**). When stratified by patient status, ICU patients developed significantly higher IgM, IgG, and IgA antibody titers than outpatients (**Fig. 2E**). Testing of the 494 remnant plasma samples from inpatients and outpatients for IgM, IgG and IgA specific for the S1 domain of SARS-CoV-2 spike showed time courses for antibody increases and declines that were very similar to those seen for RBD (**fig. S2 and table S1**). In contrast, anti-SARS-CoV-2 N antibody responses varied from those seen for spike domain antigens, particularly for IgM, which showed a strikingly low positivity rate for both outpatient and inpatient samples (**fig. S3 and table S1**).

In addition to measuring viral spike RBD-specific antibodies, we used a new competition immunoassay to test whether patient plasmas had the ability to block the binding of human ACE2 protein to the RBD. ICU patients developed significantly higher levels of ACE2-RBD blocking antibodies than outpatients (**Fig. 2A, D, and E**).

**Fig. 2.**
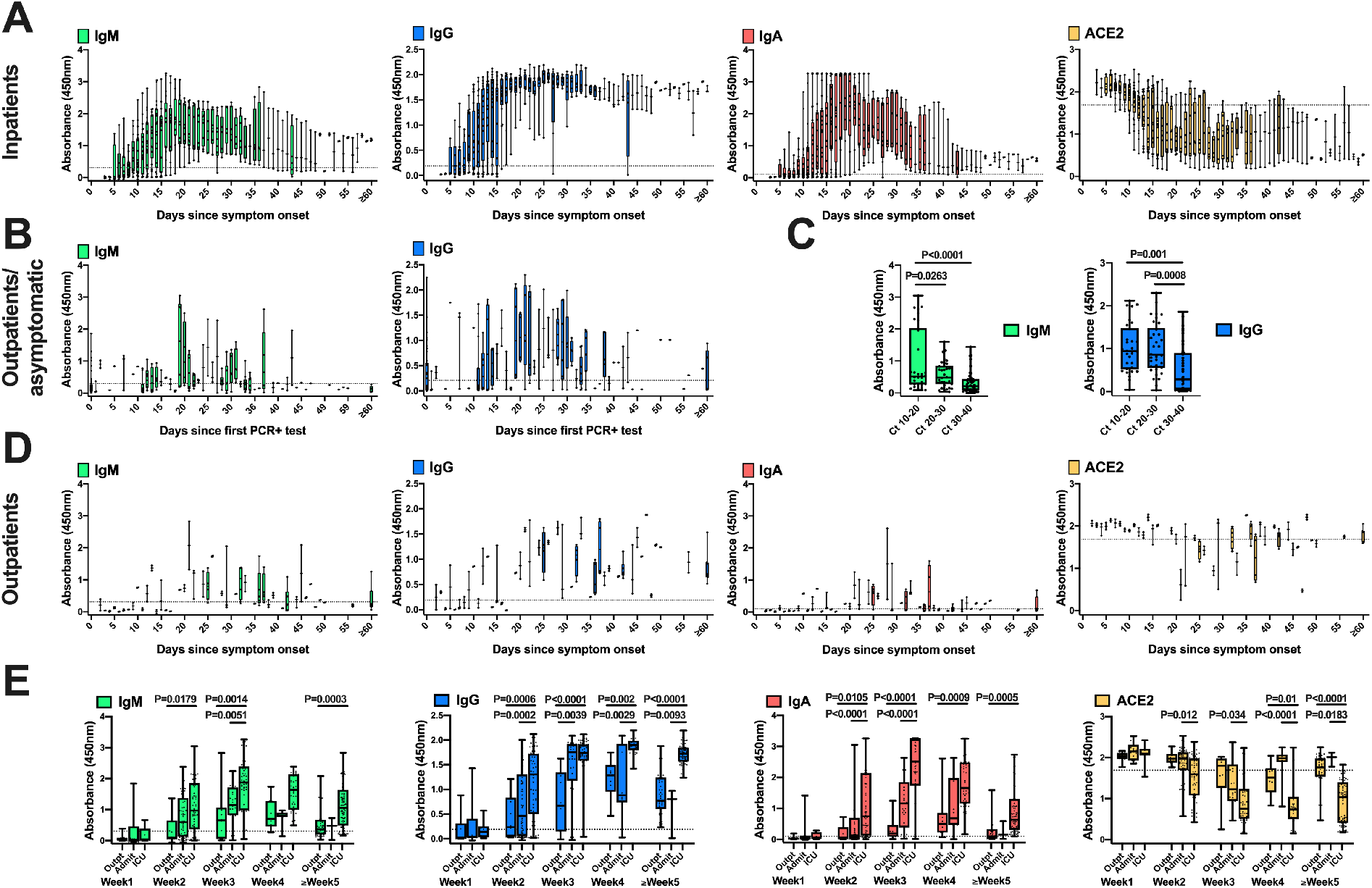
Development of anti-SARS-CoV-2 spike RBD antibody titers in SARS-CoV-2 PCR+ individuals over time. 396 plasma samples collected from 40 COVID-19 inpatients at different timepoints after symptom onset were tested by ELISA for the presence of virus-specific IgM, IgG, and IgA antibodies and for antibodies blocking binding of ACE2 to RBD. Note that the ACE2 blocking assay results are plotted as the amount of ACE2 detected binding to RBD, with lower values indicating greater blocking by patient antibodies (**A**). Initial diagnostic anti-SARS-CoV-2 RBD IgM and IgG ELISA results for 149 outpatients and asymptomatic SARS-CoV-2 rRT-PCR positive individuals are shown for sample timepoints relative to the day after individuals were first diagnosed by SARS-CoV-2 rRT-PCR. Patients who had their first rRT-PCR and serology test done on the same day are shown at x = 0 (**B**). Results for initial anti-RBD IgM and IgG ELISA testing of the 149 individuals are presented separately for subjects with low (Ct 10–20), middle (Ct 20–30) and high (Ct 30–40) diagnostic SARS-CoV-2 rRT-PCR cycle threshold (Ct) (**C**). SARS-CoV-2 RBD IgM, IgG, and IgA, and RBD-ACE2 blocking results for 84 outpatients from whom remnant plasma and date of symptom onset were available (**D**). ELISA data stratified by the 84 outpatients (Outpt), 25 hospitalized patients (Admit), and 15 patients treated in the ICU during hospitalization, for weeks 1 to 5 post-onset of symptoms are presented in **E**. In all panels, box-whisker ELISA OD_450_ plots illustrate the interquartile range as the box and the minimum and maximum values as the ends of the whiskers. The dotted lines denote the assay cutoff for seroconversion. ELISA measurements were performed in duplicate for each sample and mean ELISA OD_450_ values are shown. Comparisons between groups were by one-way ANOVA.

### The development of anti-RBD antibodies and blocking of ACE2 interaction is associated with resolution of RNAemia

Viral RNAemia is detected in up to a third of COVID-19 patients, most often in patients with severe disease *(3, 19)*. EDTA plasma was available from all inpatients for SARS-CoV-2 rRT-PCR testing. Reduction in detectable viral RNAemia was found to coincide with the appearance of plasma antibodies (**Fig. 3A** upper panels, **fig. S4**). Increases in titers of IgM, IgG and IgA were each significantly correlated with decreases in RNAemia (correlation coefficients of –0.28 for IgM, –0.34 for IgG, and –0.51 for IgA, p<0.001 for each). Further, the slopes of the increases in antibody titers and decreases in detectable RNAemia were significantly correlated. ACE2-RBD blocking antibodies were present in the majority of inpatients (68%), but showed a variable time of appearance, and extent of blocking achieved, between different individuals (**Fig. 3A** lower panels, **fig. S4**). ACE2 binding to RBD was negatively correlated with the increases in

IgM, IgG, and IgA RBD-specific antibodies (correlation coefficients of –0.5 for IgM, –0.67 for IgG, and –0.7 for IgA, p< 0.001 for each). Decreases in ACE2 binding to RBD were correlated with decreases in RNAemia (correlation coefficient 0.48, p< 0.001). The similar time course of appearance of RBD-specific IgM, IgG and IgA limited our ability to distinguish the ACE2-RBD blocking activity for each of these isotypes. Only 40% of the outpatient samples had detectable ACE2-RBD blocking activity, which was highly correlated with the presence (**Fig. 3B**) or absence (**Fig. 3C**) of anti-RBD Ig in the samples.

**Fig. 3.**
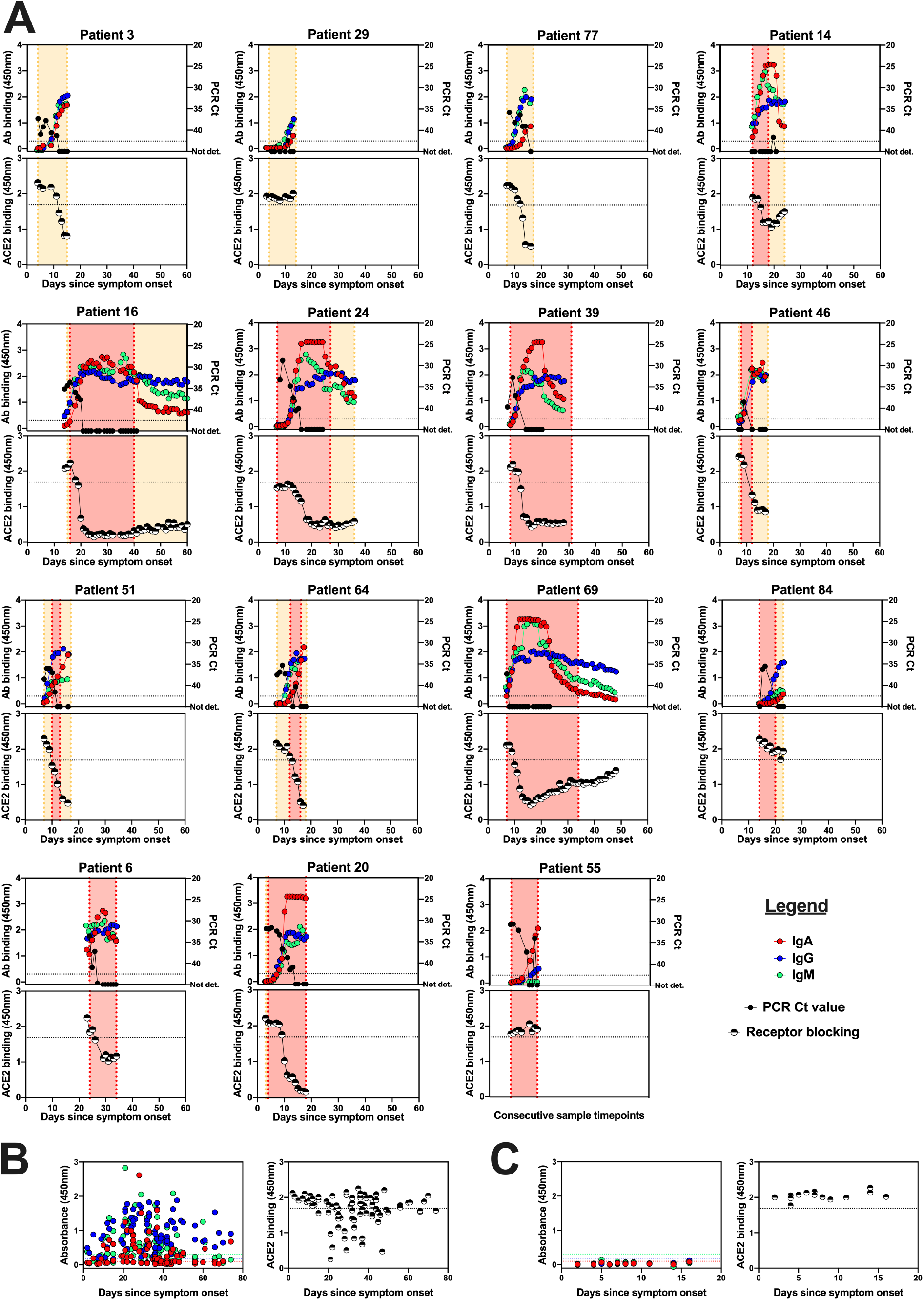
Anti-SARS-CoV-2 RBD antibodies and blocking of ACE2-RBD binding is correlated with a decrease in viral RNAemia. Plasma samples from COVID-19 inpatients were tested for the presence of SARS-CoV-2 RNA. Selected plots (shown for those 15 patients with more than 5 available ELISA and rRT-PCR measurements (with at least one being positive) illustrate ELISA OD (left y-axis) and rRT-PCR (right y-axis) data (upper panels). The dotted line denotes the cutoff for seroconversion for all isotypes. For each patient, we also tested if antibodies in plasma samples were capable of preventing binding of ACE2 to RBD in a competition ELISA (lower panels). Orange shading indicates time admitted in the Stanford hospital, red shading represents the timeframe patients were treated in the ICU (**A**). Patients 6, 20, and 55 died of COVID-19. The x-axis for Patient 55 shows consecutive sample timepoints because the onset of symptoms was unavailable for that patient. All plots for remaining patients are presented in **fig. S4**. Anti-SARS-CoV-2 RBD and competition ELISA results for outpatients, who had detectable anti-RBD Ig responses (**B**) or no detectable anti-RBD Ig responses (**C**) are shown.

### IgG binding breadth for SARS-CoV and SARS-CoV-2 RBD is common in COVID-19 patients with severe disease

Most monoclonal antibodies targeting SARS-CoV RBD fail to bind SARS-CoV-2 RBD, indicating distinct antigenicity despite sequence and structural similarity of the two proteins *(20*, *21)*. Analysis of the 494 plasma samples from inpatients (**Fig. 4A**) and outpatients (**Fig. 4B**) for the presence of RBD-specific SARS-CoV/SARS-CoV-2 cross-reactive IgG antibody titers revealed a lack of antibodies recognizing the SARS-CoV RBD in most of the outpatient plasmas.

In contrast, 9 of 15 ICU patients and 3 of the 25 non-ICU inpatients developed cross-reactive IgG titers during the course of their infection. Interestingly, the time course of anti-SARS-CoV RBD positivity in serial samples from individual patients did not always mirror anti-SARS-CoV-2 RBD IgG responses, which is particularly evident from the sharp peaks of anti-SARS-CoV levels in samples from patients 6 and 69 as opposed to the more persistent anti-SARS-CoV-2 RBD IgG in those patients (**Fig. 3A**) as well as from the absence of cross-reactive IgG responses in other ICU patients with high titers of anti-SARS-CoV-2 RBD IgG, such as patients 45, 46, and 51 (**Fig. 3A and fig. S4**). These responses likely represent limited clonal or oligoclonal B cell responses within the overall polyclonal anti-SARS-CoV-2 serological response, and suggest that severely ill COVID-19 patients not only produce higher levels, but distinct varieties of SARS-CoV-2 antibodies.

**Fig. 4.**
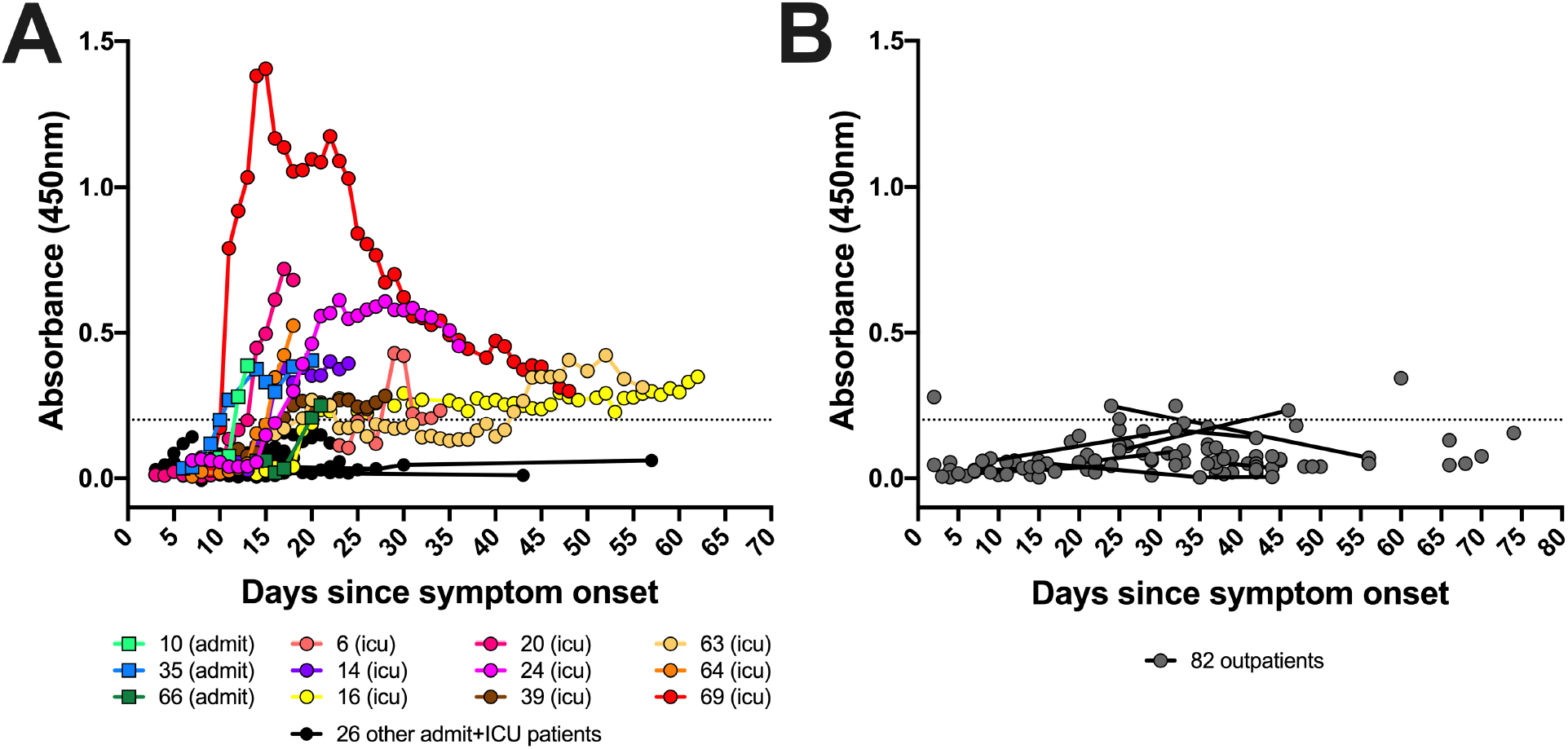
COVID-19 inpatients develop anti-SARS-CoV/SARS-CoV-2 spike RBD cross-reactive IgG responses. The 494 plasma samples from inpatients (**A**) and outpatients (**B**) were analyzed for the presence of anti-SARS-CoV spike RBD-binding IgG antibodies. The dotted line denotes the cutoff value for positivity. Squared symbols and dots represent samples from admitted non-ICU and ICU patients, respectively. Assays were performed in duplicate and mean OD values are shown.

## Discussion

Key clinical questions in the ongoing COVID-19 pandemic are the extent to which patient antibody responses may be protective against reinfection, and the duration of individuals’ serological responses. As SARS-CoV-2 vaccine candidates enter clinical trials, comparison of vaccine-induced immune responses to those stimulated by viral infection will be important for understanding immunological correlates of protection.

Here, we have analyzed serological responses to SARS-CoV-2 in 210 individuals ranging from ICU patients to asymptomatic individuals, with detailed analysis of antibody responses in 124 patients using a panel of SARS-CoV-2 antigens, and a novel assay testing the ability of RBD-specific antibodies to block binding of human ACE2. The appearance of IgM, IgG, and IgA antibodies, and the development of blocking activity preventing ACE2-RBD interaction were strongly correlated with each other, and were most prominent in severely ill COVID-19 patients. Nucleocapsid-specific responses differed from spike RBD or S1 antigen responses primarily in the low levels of IgM elicited by this antigen. RBD-specific antibody responses were tightly correlated with decreases in viral RNAemia, consistent with the humoral immune response acting to remove virus from circulation, and presumably from other sites in the body.

In contrast to some prior studies reporting detection of anti-SARS-CoV-2 IgM at earlier timepoints post-onset of symptoms compared to IgG antibody responses *(13, 22)*, we found no significant difference in the timing of detectable anti-SARS-CoV-2 RBD IgM and IgG in our patient group, as noted in more recent reports *(23)*. The timing of the onset of IgG responses and the finding that severely ill COVID-19 patients who required ICU care developed high anti-RBD antibody titers indicates that delayed or impaired production of virus-specific antibodies relative to the onset of symptoms does not explain differences in disease severity. More robust antibody responses in patients with severe as opposed to mild infection has also been reported for other coronavirus infections *(16–18)*. Notably, the IgG responses of inpatients, but not the IgM or IgA responses, were sustained at high levels for at least two months, although not all patients were observed for this length of time.

We evaluated patient plasma with a newly developed ACE2 competition ELISA to detect antibodies that can block viral RBD interaction with the host cell receptor. This assay is highly scalable and suitable for population screening as a potential surrogate for cell culture-based virus neutralization testing. We find that antibodies blocking binding of ACE2 to RBD appear in the majority of inpatients during the course of their infection. Plasma from some patients, including patient 16 (who required ICU care for an extended period of time) and patient 20 (who died of COVID-19 during the study period) contained antibodies with high blocking capacity, indicating that these antibodies, at the time that they developed in these patients, were not sufficient to prevent a deterioration of the patients’ health. Patients 6 and 55, who died of COVID-19 during the study period, did not reach full blocking capacity of ACE2-RBD binding, suggesting that the quality of their humoral responses could have contributed to their deaths, but a counter-example is seen in patient 14, who was discharged from ICU care relatively quickly despite having similarly incomplete ACE2-RBD blocking activity.

In contrast to the inpatients in our study, SARS-CoV-2-infected outpatients or asymptomatic individuals showed weaker plasma antibody responses of shorter duration, with a peak of IgG levels at approximately one month after diagnostic rRT-PCR testing, followed by a relatively rapid decline. One implication of these results is that seroprevalence studies may, over time, underestimate the proportion of the investigated population which has been previously infected with SARS-CoV-2.

To further investigate the fine specificity of patient antibody responses we tested patient plasmas for breadth of IgG binding to both SARS-CoV-2 RBD and the RBD from SARS-CoV, the causative agent of the SARS epidemic in 2003. A recent report of COVID-19 patients with unspecified disease severity at up to 22 days post-onset of symptoms found that cross-reactive antibody responses to the SARS-CoV spike ectodomain were more common than responses to the SARS-CoV RBD, and that only one of 15 patients showed cross-neutralization of SARSCoV *(24)*. Our data demonstrate that it is almost entirely in patients with severe COVID-19 illness that antibodies with breadth of binding including SARS-CoV are found, suggesting that these patients generate antibodies to distinct epitope subsets or a greater diversity of epitopes, in addition to producing higher antibody levels. We observed highly variable time courses of these cross-reactive antibodies, showing a transient appearance in some patients, and a slow increase in others. These responses to specific cross-reactive epitopes may reveal the dynamics of the frequency, extent of plasma cell differentiation, and survival of individual B cell clones amid each patient’s polyclonal response. It is likely that B cell clonal responses to other epitopes may also show variation between individuals. Given that the subgenus *Sarbecovirus* has already given rise to two coronavirus infection outbreaks in humans in the past two decades, it would be appealing if vaccination efforts could attempt to provide broad protection against this group of viruses. Our data suggest that novel vaccine strategies may be required to stimulate responses to rarely-targeted cross-reactive neutralizing epitopes.

Most outpatient plasma samples showed little ACE2 blocking capacity, although there were a few individuals that were exceptions to this pattern. ACE2 blocking activity showed a similar time course to total RBD-specific IgG antibody levels, decreasing at later time points. Recent reports find relatively low titers of neutralizing antibodies for SARS-CoV-2 or pseudotyped viruses expressing SARS-CoV-2 spike in many mildly ill to asymptomatic individuals *(25, 26)*. We find that plasma antibody responses to SARS-CoV-2 are generally of short duration following asymptomatic or mild infection, but this does not necessarily indicate that all immunity will be lost. It is possible that local antibody production in the airways could help prevent or impair SARS-CoV-2-infection upon reexposure *(27)*. Even if serum antibodies have waned to undetectable levels, memory B and T cells stimulated by infection could provide a faster or more effective response in future. Clinical trial results from patients with known reexposure to SARS-CoV-2 after recovery from initial infection will be needed to determine which serological or other immunological assays provide the most accurate correlates of protection from reinfection.

## Materials and Methods

### Study design and participants

The objective of this study was to investigate correlations between humoral immune responses to SARS-CoV-2, antibodies blocking binding of RBD to the human ACE2 receptor, and viral RNAemia in different COVID-19 patient groups and individual patients.

On March 4, 2020, the Stanford Health Care Clinical Virology Laboratory began rRT-PCR testing on nasopharyngeal specimens from suspected COVID-19 patients using a laboratory-developed SARS-CoV-2 rRT-PCR assay *(28, 29)*. For this study, we included specimens from patients with rRT-PCR-confirmed SARS-CoV-2 infection who reported with symptoms of COVID-19 to Stanford Healthcare-associated clinical sites between March 4, 2020 and April 8, 2020; and specimens from rRT-PCR positive outpatients and asymptomatic individuals identified between April 8, 2020 and May 26, 2020 through occupational health screening including rRT-PCR and serology testing for anti-RBD IgM/G at Stanford Clinical Laboratories.

### Sample and data collection

Venipuncture blood samples collected in sodium heparin- or K_2_EDTA-coated vacutainers were used for serology testing and real-time PCR detection of RNAemia, respectively. After centrifugation for collection of plasma, samples were stored at –80°C.

Retrospective chart review was performed on 124 patients with remnant plasma samples for detailed serological analyses. Collected data included age, gender, date of symptom onset, admission to hospital, admission to ICU, and date and Ct for the diagnostic nasopharyngeal swab rRT-PCR test result. For inpatients we also recorded presence of underlying comorbidities, clinical symptoms, and mortality. Collected data for the 86 patients, from whom no remnant plasma samples were available were initial IgM and IgG Clinical Laboratory serology test result, and date and Ct for the diagnostic nasopharyngeal swab rRT-PCR test result.

### Production of SARS-CoV and SARS-CoV-2 proteins and ACE2-mFc

The SARS-CoV and SARS-CoV-2 RBD proteins were expressed in Expi293F cells and purified using Nickel-NTA resin and size exclusion chromatography. The SARS-CoV construct (RBDHis_pTT5, GenBank AAP13441.1) was synthesized commercially by Twist Bioscience (San Francisco, CA); the SARS-CoV-2 construct (RBD-His_pCAGGS, GenBank MN908947.3) was kindly provided by Dr. Florian Krammer *(30)*. SARS-CoV-2 S1 (spike residues 1–682) and ACE2-mFc, expressed in HEK293 cells, and the N protein, expressed in *E.coli* were produced by the CRO Atum. Soluble human ACE2 fused to a mouse Fc tag was constructed by synthesizing a gene encoding ACE2 (residues 1–615) joined by a (G_4_S)x2 linker to a mouse IgG2a Fc, and placed under control of a CMV promoter by cloning into a mammalian expression plasmid.

### ELISA to detect anti-SARS-CoV-2 Spike RBD antibodies in plasma samples

The ELISA procedure in this study was modified from a protocol published by Stadlbauer *(30)*.96-well Corning Costar high binding plates (catalog no. 9018, Thermo Fisher) were coated with SARS-CoV RBD, SARS-CoV-2 RBD, S1, or nucleocapsid protein in phosphate-buffered saline (PBS) at a concentration of 0.1 µg per well (0.025 µg per well for the nucleocapsid IgG assay) overnight at 4°C. On the next day, wells were washed 3x with PBS – 0.1% Tween 20 (PBS-T) and blocked with PBS-T containing 3% non-fat milk powder for 1 hour at room temperature (RT). Wells were then incubated with plasma samples from COVID-19 patients at a dilution of 1:100 in PBS-T containing 1% non-fat milk for 1 hour at 37°C. Two negative and two positive plasma pool wells and two blank wells incubated with PBS-T containing 1% non-fat milk powder were included on each plate. After washing 3x with PBS-T, horseradish peroxidase conjugated goat anti-human IgG (g-chain specific, catalog no. 62–8420, Thermo Fisher, 1:6’000 dilution), IgM (µ-chain specific, catalog no. A6907, Sigma, 1:6’000 dilution), or IgA (a-chain specific, catalog no. P0216, Agilent, 1:5’000 dilution) in PBS-T containing 1% non-fat milk was added and incubated for 1 hour at RT. Wells were washed 3x with PBS-T and dried by vigorous tapping of plates on paper towels. 3,3’,5,5’-Tetramethylbenzidine (TMB) substrate solution was added and the reaction was stopped after 12 minutes by addition of 0.16 M sulfuric acid. The optical density (OD) at 450 nanometers was measured with an EMax Plus microplate reader(Molecular Devices, San Jose, CA); values for blank wells were subtracted from values obtained for plasma samples. The cutoff value for seroconversion was calculated by adding 3 standard deviations to mean ELISA ODs of 90 historical negative control samples from healthy blood donors (collected before the pandemic for an unrelated seroprevalence study) obtained by testing in all protein/isotype assays. Additional details for the manual and clinical lab instrument ELISA assay setup are provided in **figs. S5 and S6**.

### Competition ELISA to detect antibodies blocking binding of ACE2 to RBD

All competition ELISA steps were carried out on the same day at RT. 96-well Corning Costar high binding plates (Thermo Fisher: cat. 9018) were coated with SARS-CoV-2 Spike RBD protein in PBS at a concentration of 0.1 µg per well for 3 hours (or overnight at 4°C). Wells were washed 3x with PBS-T and blocked with PBS-T containing 3% non-fat milk powder for 90 minutes. Wells were then incubated with plasma samples from COVID-19 patients at a dilution of 1:50 in PBS-T containing 1% non-fat milk for 105 minutes. A negative and a positive plasma pool and two blank wells incubated with PBS-T containing 1% non-fat milk were included on each plate. ACE2-mFc diluted to 0.2 µg/ml in 1% non-fat milk powder was added without washing steps and incubated for an additional 45 minutes. After washing 3x with PBS-T, horseradish peroxidase conjugated goat anti-mouse IgG (Fc specific, catalog no. 31439, Invitrogen, 1:20’000 dilution) in PBS-T containing 1% non-fat milk was added and incubated for 45 minutes. Wells were washed 3x with PBS-T and dried by vigorous tapping of plates on paper towels. TMB substrate solution was added and the reaction was stopped after 12 minutes by addition of 0.16 M sulfuric acid. The OD at 450 nanometers was measured with an EMax Plus microplate reader (Molecular Devices, San Jose, CA). Additional details for competition ELISA assay setup are provided in **fig. S7**.

### Real-time PCR to detect SARS-CoV-2 RNA in plasma

A volume of 400 µL of EDTA-anticoagulated plasma was extracted by Qiagen EZ1 Virus Mini Kit v2.0 (Qiagen Germantown, MD). Molecular testing for the presence of SARS-CoV-2 RNA in plasma was performed with a modification of a published rRT-PCR assay targeting the envelope (*E*) gene *(28, 29)*. The standard Ct values of positive tests with this assay range from Ct <20 to 45 cycles. Testing of plasma samples with a Ct value of 40 or greater was repeated to ensure reproducibility of the positive result. As viral culture was not performed as part of this study, presence of SARS-CoV-2 in tested plasma was defined as RNAemia.

### Statistics

ELISA data were analyzed using GraphPad Prism version 8.0 (GraphPad Software, San Diego, California, USA). All statistical analyses, including correlations, t-tests, analysis of variance and least squares slopes computations were carried out using the R statistical package version 3.6.1 *(31)*. For statistical assessment of differences between patient groups we adjusted for the fact that the median time of sample collection after onset of symptoms was different for outpatients (median of 30 days), admitted patients not treated in the ICU (median of 12 days) and patients treated in the ICU (median of 22 days) by comparing results in the same timeframe (i.e. results within week 1, week 2, week 3, week 4, and week 5 after symptom onset).

Correlation between antibody OD_450_ values, RNAemia, and ACE2-RBD blocking assay OD_450_ values were calculated with the R cor function.*(31)* The p-values for the correlation between the slopes of anti-RBD antibodies and RNAemia were estimated using 1000 permutations of the RNAemia values within each patient time course. The p-values for the correlation between the slopes of anti-RBD antibodies and ACE2 were estimated using 1000 permutations of the ACE2 values within each patient time course. The p-values for the difference in slopes of anti-RBD antibody decreases (after they peaked) between outpatients/asymptomatic individuals and inpatients were estimated using 1000 permutations of the group labels for each patient. Slopes were computed using linear regression analysis. A p-value of <0.05 was considered significant.

This study was approved by the Stanford University Institutional Review Board (protocol # 48973).

## Data Availability

All data is available in the main text or the supplementary materials.

## Supplementary Materials

Fig. S1. Study design and participant overview.

Fig. S2. Development of anti-SARS-CoV-2 spike S1 antibody titers in COVID-19 patients.

Fig. S3. Development of anti-SARS-CoV-2 N antibody titers in COVID-19 patients.

Fig. S4. Anti-SARS-CoV-2 RBD antibodies are correlated with a decrease in viral RNAemia.

Fig. S5. Checkerboard titration for serological RBD ELISA.

Fig. S6. Validation of the Clinical Lab anti-RBD IgM/G ELISA.

Fig. S7. Checkerboard titration for receptor blocking ELISA

Table S1. Inpatient seropositivity.

Table S2. Outpatient and asymptomatic individuals’ seropositivity.

## Acknowledgments

The authors thank Dana Anderson for helpful advice about protein production, and the ATUM Bio team for optimization and production of antigens.

## Author contributions

K.R, O.F.W., B.A.S., A.E.P., B.A.P., S.D.B. conceptualized and designed the study. K.R., O.F.W., A.E.P., C.A.H., M.K.S., J.M. performed the experiments. B.A.S., M.K.S., T.D.P., A.R., A.J.R., C.A.B., J.R.C., K.C.N., B.A.P., J.N., S.G., N.H.S. collected data and/or contributed samples/reagents or EHR processing methods. C.A.H., J.M., T.S.J., J.L.Z., T.T.W., P.S.K., provided intellectual contributions throughout the study. R.T. performed statistical analyses. K.R., O.F.W., B.A.P., C.A.H., S.G., R.T., S.D.B. analyzed the data. K.R., S.D.B. wrote the manuscript. All authors edited and approved the manuscript.

## Data and materials availability

All data is available in the main text or the supplementary materials.

